# SARS-CoV-2 seroprevalence and living conditions in Bamako (Mali): a cross-sectional multistage household survey after the first epidemic wave, 2020

**DOI:** 10.1101/2022.06.03.22275924

**Authors:** Mady Cissoko, Jordi Landier, Bourema Kouriba, Abdoul Karim Sangare, Abdoulaye Katile, Abdoulaye Djimdé, Ibrahima Berthé, Siriman Traoré, Ismaïla Thera, Hadiata Maiga, Elisabeth Sogodogo, Karyn Coulibaly, Abdoulaye Guindo, Ousmane Dembelé, Souleymane Sanogo, Zoumana Doumbia, Charles Dara, Mathias Altmann, Emmanuel Bonnet, Hubert Balique, Luis Sagaon-Teyssier, Laurent Vidal, Issaka Sagara, Marc-Karim Bendiane, Jean Gaudart

## Abstract

**Context:** In low-income settings where access to biological diagnosis is limited, data on the spread of the COVID-19 epidemic are scarce. In September 2020, after the first COVID-19 wave, Mali reported 3,086 confirmed cases and 130 deaths. Most reports originated form Bamako, the capital city, with 1,532 reported cases and 81 deaths for an estimated 2.42 million population. This observed prevalence of 0.06% appeared very low. Our objective was to estimate SARS-CoV-2 infection among inhabitants of Bamako, after the first epidemic wave. We also assessed demographic, social and living conditions, health behaviors and knowledge associated with SARS-CoV-2 seropositivity.

**Material and methods:** We conducted a cross-sectional multistage cluster household survey in commune VI, which reported, September 2020, 30% (n=466) of the total cases reported at Bamako. We measured serological status by detection of SARS-CoV-2 spike protein Antibodies in venous blood sampled after informed consent. We documented housing conditions and individual health behaviors through KABP questionnaires among participants aged 12 years and older. We estimated the number of SARS-CoV-2 infections and deaths in the total population of Bamako using the age and sex distributions of SARS-CoV-2 seroprevalence. A logistic generalized additive multilevel model was performed to estimate household conditions and demographic factors associated with seropositivity.

**Results:** We recruited 1,526 inhabitants in the 3 investigated areas (commune VI, Bamako) belonging to the 306 sampled households. We obtained 1,327 serological results, 220 household questionnaires and collected KABP answers for 962 participants. The prevalence of SARS-CoV-2 seropositivity was 16.4% after adjusting on the population structure. This suggested that ∼400,000 cases and ∼ 2,000 deaths could have occurred of which only 0.4% of cases and 5% of deaths were officially reported. KABP analyses suggested strong agreement with washing hands but lower acceptability of movement restrictions (lockdown or curfew), and limited mask wearing.

**Conclusion:** In spite of limited numbers of reported cases, the first wave of SARS-CoV-2 spread broadly in Bamako. Expected fatalities remained limited largely due to the population age structure and the low prevalence of comorbidities. This highlight the difficulty of developing epidemic control strategies when screening test are not available or not used, even more when the transmission modalities are not well known by the population. Targeted policies based on health education prevention have to be implemented to improve the COVID-19 risk perception among the local population and fight to false knowledge and beliefs.

## Background

COVID-19 disease, due to the Severe Acute Respiratory Syndrome Coronavirus 2 (SARS-CoV-2), which emerged at the end of 2019 in Wuhan, China, has spread rapidly around the world and was declared as “pandemic” on 11 March 2020 by the World Health Organization (WHO) [1]. Despite setting up public health policies appropriated to this pandemic situation, such as lockdown, quarantine and curfew, the virus continues to circulate [2, 3] The WHO African Region reported the least number of affected people since the pandemic began. Indeed, in many resource-limited settings, biological confirmation was only available in tertiary medical facilities and has been reserved for symptomatic patients (mostly severe) and/or travelers, the various national policies requiring a negative test for travel. As a result, the number of people exposed to the virus in Sub-Saharan Africa is still largely unknown [1].

After the first reported case on March 25^th^ 2020 (coming from France on March 12^th^), Mali has recorded, 6 months later (at the time of the survey), 3,086 cases of SARS-CoV-2 diagnosed by RT-PCR, i.e. an incidence rate of 0.015% for the whole country. Spread over 38 health districts (among 75), they led 130 reported deaths, *i*.*e*. a case fatality rate of 4.2%[4]. Among the recorded cases at M6 (September 2020), ∼50% were reported in the district of Bamako *i*.*e*. 1,532 reported cases, for a population of at least 2.42 million inhabitants. The most affected area was the Commune VI with 466 reported cases and 27 associated deaths. The second largest number of recorded cases was reported in the region of Timbuktu, with 572 confirmed cases at M6 [4]. Given the limited access to diagnosis and care, and in the absence of a reliable syndromic surveillance, the low number of reported cases did not allow to assess accurately the epidemic situation. In this context, serological surveys represent an important tool to assess the extent of the exposure to SARS-CoV-2 in the general population. A single survey provides a snapshot of the extent of the virus spread at a given time point, and informs on vulnerable population groups, on the denominators used to calculate infection fatality rate or hospitalization rates [5]. In Mali, a multi-site study including a peri-urban area of the capital city Bamako demonstrated a sharp increase in seroprevalence between a survey conducted after the first wave of clinical cases (August 2020) and a survey conducted during the decrease of the second wave (January 21), identifying geographical location and age as associated factors [6]. Indeed, Sagara *et al*. reported in the peri-urban area of Sotuba a crude seroprevalence of 13.1 % (n=587) after the first wave. In the capital city of Kinshasa, Nkuba *et al*. reported a similar result with a seroprevalence of 16.6% (n=1233) [7]. Seroprevalence is also essential to assess the level of herd immunity that has been developed, which determines the risk of the following epidemic waves, their potential severity and their potential impact on the healthcare system. Measuring immunity could also help develop response strategies including priority strains for vaccination or targeted awareness campaigns. In the settings where mortality and hospitalization statistics are not readily available, approximating the number of infections by age groups and by gender was also important to estimate the order of magnitude for expected infection fatality rates and compare it to reported COVID-19 deaths [8]. In addition, better access to information on epidemiological trends, social factors associated, health and protective behaviors, as well as attitudes and beliefs, was needed to design control strategies and strengthen information and awareness campaigns. The aim of this study was to estimate the seroprevalence of SARS-CoV-2 in the population of the most populated and affected commune of Bamako, after the first epidemic wave. We also assessed demographic, social and living conditions; health behaviors; and knowledge associated with SARS-CoV-2 seropositivity.

## Material and methods

### Study design and sample size calculation

In accordance with the WHO guidelines protocol for age-stratified population-based sero-epidemiological surveys for COVID-19 infection, a cross-sectional household survey was conducted [8] in the 3 most affected and populated neighborhoods of Bamako’s commune VI: Faladié, Banakabougou, and Yirimadjo (Figure 1), September 2020. At the time of the protocol (July 2020), the number of cases reported was 38, 29, and 40 respectively for these neighborhoods, representing 0.07 cases/ 100 inhabitants, and 54% of the total reported cases in Commune VI.

**Figure 1:**
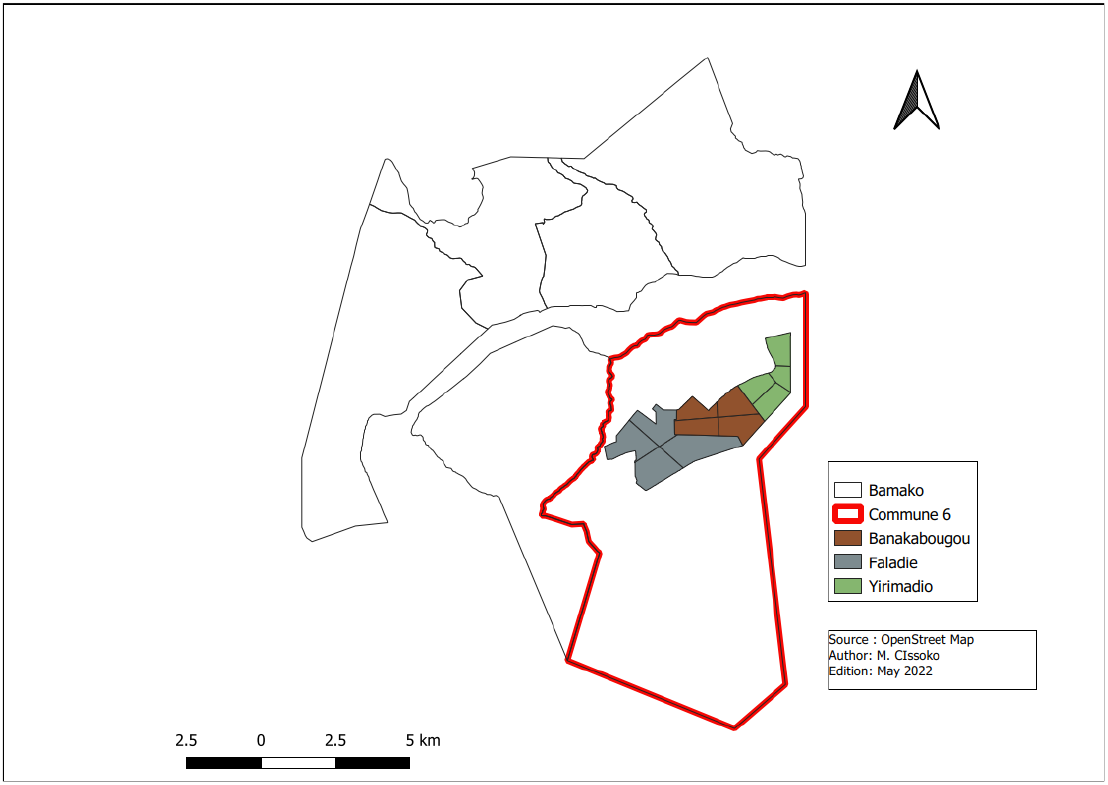
Map of Bamako showing the location of the 3 investigated neighborhoods within the Commune VI (in red).

The sample size was calculated assuming an expected prevalence of COVID-19 infection of 0.07 cases/ 100 inhabitants, within the population. Based on this assumption, a sample size of 1300 persons was estimated, with a precision of 2% and a confidence interval of 95%. Considering 15% loss, 1500 people was expected to be included. A multi-stage cluster sampling method covering all age ≥1 groups of the population was performed [9]. In the first stage, the sample size to be recruited per district was proportional to the district population sizes. In the second stage, each district was divided into different sectors (4 or more) of relatively equal sub-population size. The household survey therefore concerned each sector of each district. The first household in each sector was selected by choosing a random direction from the center of the community sector, counting the houses along that road and selecting one at random. Subsequent households were selected by visiting the closest house to the previous one. All household members in the age range willing to participate were recruited. The study was conducted among the general population aged ≥1-year-old for the seroprevalence study, and ≥12-year-old for the questionnaire survey. A housing unit was defined as a private one, such as apartment or villa or collective house (living quarter called “compound”) with its own separate entry. Common residence rules (*de jure* rules) defined household unit as group of first-degree relatives usually living in the same housing unit. This approach allowed considering Malian family structure and local housing habits to define household units.

### Individual sample and data collection

After informed consent obtained from the participants or their parents, 2mL of blood were collected from all voluntary participants by venipuncture (September 2020), to perform serological tests. Following the blood sampling, a face-to-face questionnaire was administered to collect the following demographic and sociologic factors: gender, age, history of recent travel within and outside Bamako, socio-economic level, contact with COVID-19 cases, occupation, education level, recent clinical symptoms, recent treatment, and attendance at places of worship. The questionnaire also included items relative to the knowledge about the disease, protective measures and consequences on the population health.

### Housing conditions and household data collection

The head of household was asked to answer a specific questionnaire documenting their individual characteristics (age, gender, education, profession), household structure (number and age of members) and housing conditions including housing equipment, goods, and incomes of family (auto, TV, moto, cell phone, external funding…).

### Biological analyses

The level of exposure of the population to SARS-CoV-2 was estimated by serology. Sera were separated from whole blood and stored at -80°C in cryotubes. SARS-CoV-2 specific IgM and IgG antibodies were assayed in sera by VIDAS® anti-SARS-CoV-2 IgM and anti-SARS-CoV-2 IgG kits (BioMerieux, Lyon, France) [10]. The VIDAS® anti-SARS-CoV-2 IgM and anti-SARS-CoV-2 IgG tests relied on the SARS-CoV-2 Spike protein immunoassay technique to measure the presence of antibodies in infected participants. Compared to PCR, the sensitivity of the VIDAS® tests for IgM and IgG is 90.4% and 88.6%, 8-15 days after SARS-CoV-2 infection, 100 and 96.6%, 16 days after infection, respectively. The specificity for IgM and IgG is 99.4% and 99.6%, respectively. In this context, the specificity of the tests was particularly important to ensure that the test of an un-infected participant was indeed systematically negative. Serology analyses were performed at the Charles Mérieux Infectiology Centre in Bamako, Mali. Participants were defined as SARS-CoV-2 seropositive if they presented either a positive IgG or IgM result. Individuals were defined as SARS-CoV-2 seronegative if they presented a negative IgG and IgM result, or a negative IgG and a missing IgM result. Individuals with missing IgG results were excluded from the seroprevalence analysis. The seroprevalence was estimated as the number of SARS-CoV-2 seropositive by the number of participants. The number of infections for the district of Bamako was estimated using the population of Bamako by sex and age categories. The number of deaths was estimated by using the age-and sex-specific mortality data reported early in the pandemic (February-March in China, prior to the optimization of clinical management) [11].

### KABP outcomes measures

The current at-risk practices have been measured using a four bipolar Likert Items on practices during the seven past days assessing: wearing mask when not at home, washing hands with soap, going to crowned areas during the day or the night. Concerning behavior questions, six bipolar Likert Items (from systematically/very often to never) on behavior changes since the start of the epidemic focusing on: washing hands, visiting friends and relatives, going to crowned areas, touching each other, sneezing into elbow, reducing travel. Concerning knowledge questions, a scale-score based on 13 items (True/False/Don’t know) on prevention, treatment, symptoms, and transmission of SARS-CoV-2 has been build up. At least, concerning cultural beliefs, four bipolar Likert Items (from very agreed to very disagreed) assessed opinion about the disease focusing on infection origin: a divine punishment, a spell casting, a white people illness, a way to get money for rich people.

### Data analysis

First, descriptive analyses estimated mean, prevalence and frequencies, associated with 95% confidence intervals (95%CI). Household profiles were determined by using 2 step descriptive approach [12]: first a multiple component analysis (MCA), second a Hierarchical Ascendant Classification (HAC). Based on household level variables, this approach led to determine classes according to the different household profiles. Each individual was assigned to its household profile.

Second, in order to estimate factors associated with SARS-CoV-2 seropositivity, we used logistic generalized additive multilevel models (GAMM) [13]. We analyzed the effects of age and sex at individual level, as well as household profile. Intra-household contamination was assessed as a binary variable (more than 1 positive case or not). The GAMM approach allowed also verifying the non-linear effect of continuous covariates by using spline smoothing [14]. The model included random effects for household, compound and district sector to reflect sampling structure and potential correlations between participants sharing the same living space (household nested in compound sampled in the same sector). Main statistical tests were performed using an α-probability threshold of 5%, but with Bonferroni correction for sub-group analysis.

Data analyses were performed using the SPSS software (IBM Corp. Released 2020. IBM SPSS Statistics for Windows, Version 27.0. Armonk, NY: IBM Corp) for the questionnaire data management and descriptive analyses, and the R software (version 4.0.0, R Core Team 2020. R Foundation for Statistical Computing, Vienna, Austria.) with the following specific packages: {FactoMineR}, {lme4}, {gamm4}.

### Ethics and regulations

The authorization to conduct the study was obtained on August 28^th^, 2020, from the Ministry of Health and Social Affairs of Mali (decision letter number 2020-001424-MSAS-SG). Clearance from the ethics committee of the Faculties of Medicine and Odonto-Stomatology and Pharmacy, University of Sciences, Technics and Technologies of Bamako (Mali) was obtained on August 10^th^, 2020 (clearance letter number 2020/162/CA/FMOS/FAPH). First, a community agreement was first obtained from district leaders, local religious leaders, community associations and municipal authorities after explanation and discussion about the study protocol. Second, consents and/or assents of participants or their parent/guardian were obtained. The study team administered consent in local languages, and, if the participant or parent/guardian was not literate, in the presence of a witness. Individuals from each family consented separately.

## Results

### Inclusions

A sample of 174 housing units (separate living quarter) were investigated including 2,015 inhabitants grouped in 306 identified household units. Of 2,015 inhabitants, 1,526 (75.7%) participants aged ≥1 year provided a blood sample for the seroprevalence survey and 962 participants aged ≥12 years answered the KABP survey (Table 1). Data on housing conditions were collected for 220 of the 306 household units included, *i*.*e*. 78.9% of the household members tested (n=1,204) (Figure 2).

**Table 1:** Study participants’ demographic characteristics and detailed serological results (Bamako, n=1,526, September 2020).

|  |  | n | % |
| --- | --- | --- | --- |
| Site |  |  |  |
|  | Banankabougou | 588 | 38.5 |
|  | Faladie | 300 | 19.7 |
|  | Yirimadio | 638 | 41.8 |
| **Gender** |  |  |  |
|  | Homme | 599 | 39.3 |
|  | Femme | 927 | 60.7 |
| **Age** |  |  |  |
|  | [1-10y] | 416 | 27.3 |
|  | [10-20y] | 491 | 32.2 |
|  | [20-30y] | 299 | 19.6 |
|  | [30-40y] | 144 | 9.4 |
|  | [40-60y] | 118 | 7.7 |
|  | [60-100] | 51 | 3.3 |
|  | NA | 7 | 0.5 |
| **SARS-CoV-2 statut** |  |  |  |
|  | Negative | 1100 | 72.1 |
|  | Positive | 227 | 14.9 |
|  | NA | 199 | 13.0 |
| **Detailed pos** |  |  |  |
| **IgG pos only** |  | 170 |  |
| **IgM pos only** |  | 17 |  |
| **IgG+IgM** |  | 35 |  |
| **IgG pos, IgM missing** |  | 5 |  |

**Figure 2:**
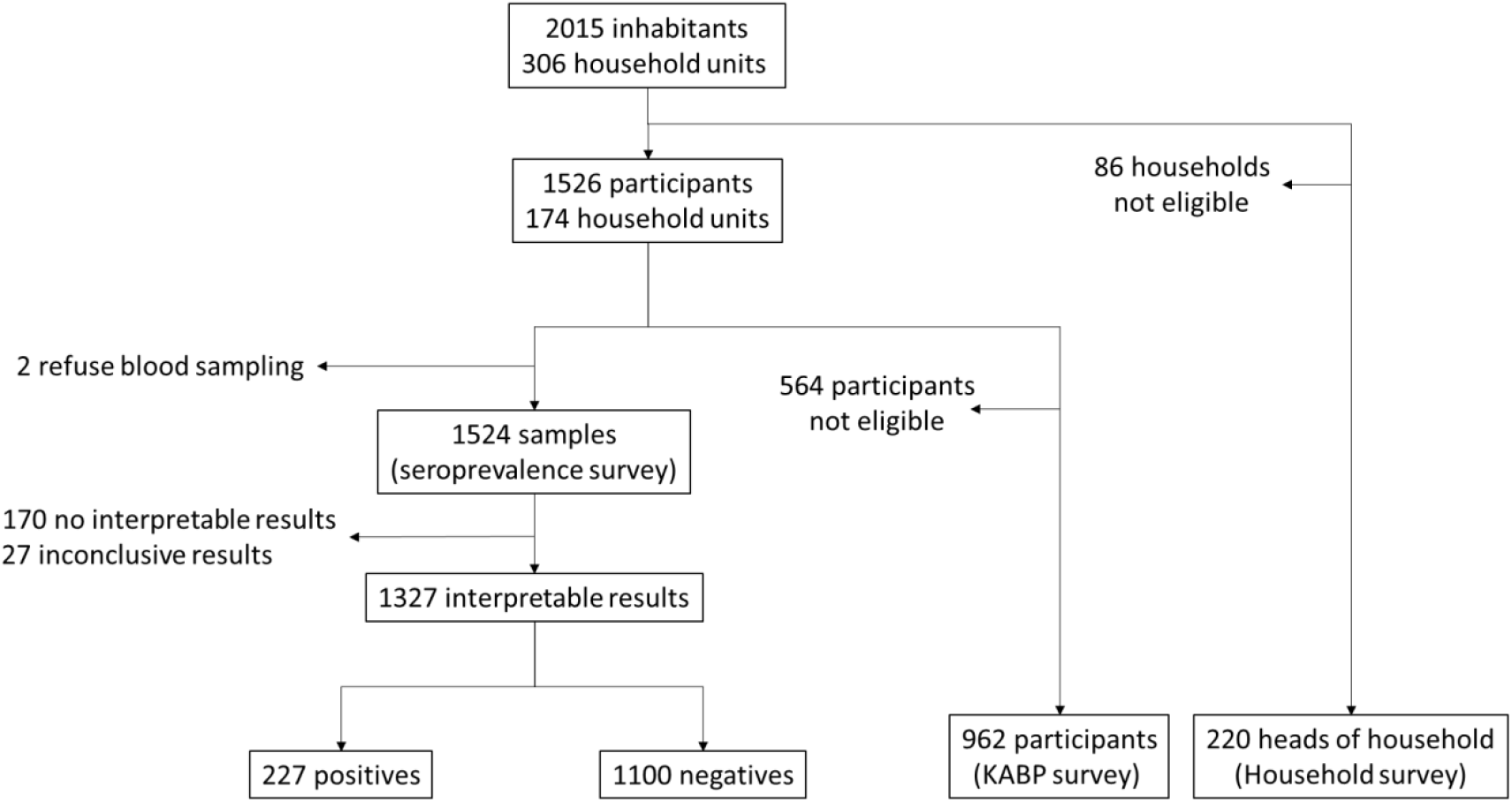
flowchart of the seroprevalence survey.

### SARS-CoV-2 seroprevalence

Out of 1,526 participants, 2 did not provide samples, 170 had no interpretable test results for both IgG and IgM, and 27 inconclusive results due to a missing IgG and negative IgM results or inversely. Overall, interpretable serological results were available for 1,327 participants, corresponding to 227 SARS-CoV-2 seropositive (by either IgG, IgM or both) and 1,100 seronegative individuals. The crude seroprevalence rate was estimated at 17.1% (95% Confidence interval (95%CI) [15.1-19.1], ranging from less than 10% to upper than 30% across genders and age groups (Figure 3).

**Figure 3:**
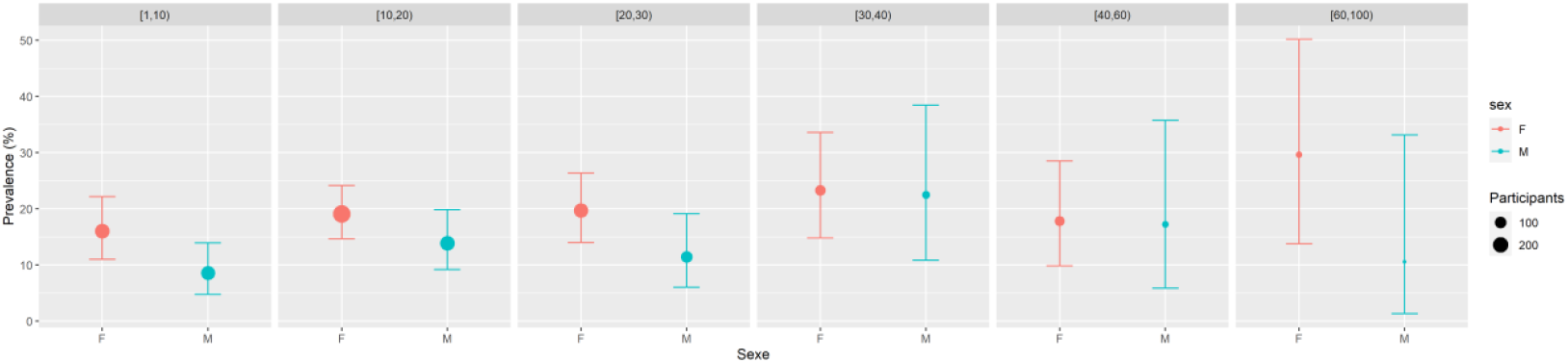
Seroprevalence by age and sex (Bamako, n=1327, September 2020).

Applying estimated prevalence, by age and sex, to the population of the district of Bamako (2.42 million inhabitants), we estimated around 400,000 the number of infections in the city between the onset of the epidemic and the time of the survey (September 2020), compared to 1,532 recorded cases for the district of Bamako. This corresponded to an adjusted prevalence of 16.4% (adjusted on the population age and sex distribution) vs an observed prevalence of 0.06%. Using the age- and sex-specific mortality data reported early in the pandemic, we roughly estimated 1,725 COVID-19 deaths occurred between the onset of the pandemic and the date of the survey, *i*.*e*. more than twenty times the 81 official reported deaths (Table 2). According to these estimates, the detection rates were low, with only 0.4% of cases and 5% of deaths reported.

**Table 2:** SARS-CoV-2 seroprevalence in the study sample, and estimated vs reported cases and deaths at Bamako city level after accounting for age population structure (Bamako, n=1,526, September 2020).

|  |  |  |
| --- | --- | --- |
| **SARS-CoV-2 serological status** |  |  |
|  | negative | 1,100 |
|  | positive | 227 |
|  |  | 17.1% [15.1-19.2] |
| **Population** |  |  |
|  | inhabitants in 2020 | 2,420,000 |
| **Prevalence** |  |  |
|  | Reported COVID cases (%) | 1,532 (0.07%) |
|  | Estimated infections (%) | 397,321 (16.4) |
| **COVID-19 associated Deaths** |  |  |
|  | Reported deaths | 81 (0.3) |
|  | Estimated deaths | 1,725 (7.1) |

### Household profile as social proxy

Among the 220 households documented, 64.6% (n=142) lived in a private house, 19.1% (n=42) shared their house with another family and 12.3% (n=27) with two others. Only 0.9% (n=2) shared their house with more than two other families (three or four).

Assessing social characteristics and housing conditions, three specific profiles have been determined related to three social dimensions (Table 3): i) Location and family structure; ii) Goods and incomes; iii) Housing conditions. The first profile selected was labelled “Poor Small Family” unit (PSF, n=62), and the second “Poor Large Family” unit (PLF, n=117). These two profiles, mainly located at Yirimadio and Banankabougou, were associated with a low level of incomes or goods, and deleterious housing conditions. The main difference between these two profiles came from the household size: 8.1% of large family (>10 members) vs 27.4% (p=0.002). The PSF profile showed also slightly (but significant) less livestock than the PLF profile (8.1% vs 12.8%, p<0.001), slightly more private toilets (24.2% vs 19.7%, p<0.001), and less rooms (14.5% vs 33.3%, p<0.001). Both profiles showed a low level of education (resp. 35.5% and 46.2% of no education), and around 50% of private house.

**Table 3:**
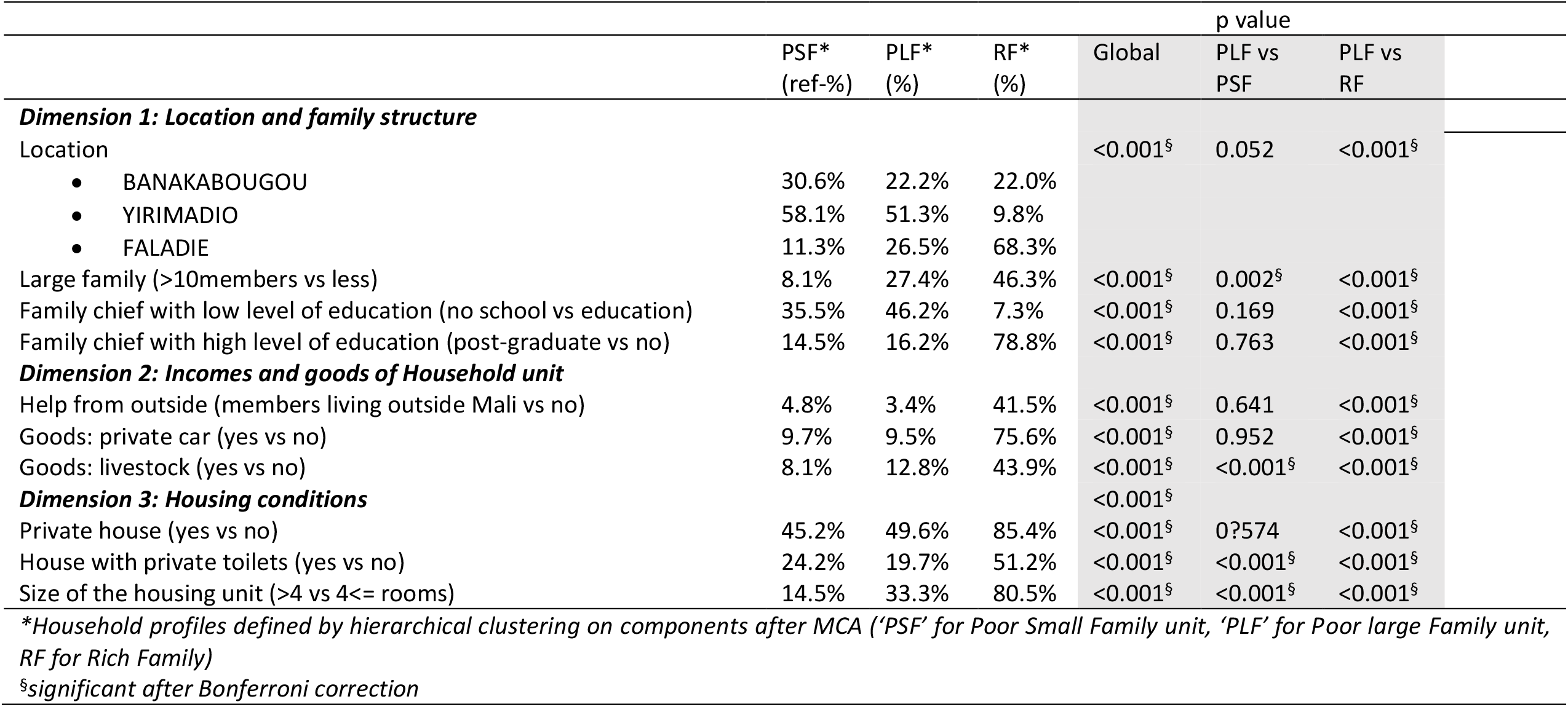
Household units’ main characteristics (Bamako, n=220, September 2020)

The third and last profile, mainly located at Faladie (68%), showed significant high level of incomes (75.6% with a private car, 41.5% having an external financial help, 43.9% having livestock) and best housing conditions (95.4% having a private house, 51.2% having private toilets, 80.5% having more than 4 rooms), and, consequently, was labelled “Rich Family” unit (RF, n=41).

### Factors associated with SARS-CoV-2 seropositivity

Factors associated with SARS-CoV-2 seropositivity were identified with a multilevel logistic regression approach (table 4) (individual, household and neighborhood levels). There were no significant differences between the three neighborhoods. Women and older age were significantly associated with increased odds of seropositivity, showing respectively adjusted Odd Ratios (aOR [CI95%]) of 1.75 [1.27;2.43] and 1.06 [1.01;1.11]. Having a positive household member was associated with an increased odd of seropositivity (aOR=1.54 [1.08;2.19]). Household corresponding to the highest socio-demographic status appeared to have increased (but not significant, p=0.06) odds of seropositivity compared to households of poor status living in (aOR=1.74 [0.99;3.07]).

**Table 4:** Factors associated with SARS-CoV-2 seropositivity in September 2020, Bamako, Mali. Unadjusted (univariate) and adjusted (multivariate) Odds-ratios (OR) after multilevel logistic regression including random effects at sector, compound and household levels.

|  |  | SARS-CoV-2<br>serology<br>n(%) / median (IQR) |  | Univariate* |  | Multivariate* |  |
| --- | --- | --- | --- | --- | --- | --- | --- |
|  |  | neg | pos | OR [CI95%] | p | aOR [CI95%] | p |
| **Sex** | **Male** | 456 (87.2) | 67 (12.8) | 1 |  | 1 |  |
|  | **Female** | 644 (80.1) | 160 (19.9) | 1.78 [1.28;2.49] | <0.001<sup>§</sup> | 1.75 [1.27;2.43] | <0.001<sup>§</sup> |
| **Age\*** | **(+1 years)** | 16 (9-25) | 18 (11-30) | 1.07 [1.02;1.12] | 0.008<sup>§</sup> | 1.06 [1.01;1.11] | 0.017<sup>§</sup> |
| **Household profile** | **Poor Small Family units** | 304 (84.9) | 54 (15.1) | 1 |  | 1 |  |
|  | **Poor Large Family units** | 456 (83.1) | 93 (16.9) | 1.08 [0.67;1.74] | 0.75 | 1.14 [0.74;1.74] | 0.56 |
|  | **Rich Family units** | 119 (75.3) | 39 (24.7) | 1.66 [0.88;3.12] | 0.12 | 1.74 [0.99;3.07] | 0.06 |
|  | **Unclassified** | 221 (84.4) | 41 (15.6) | 0.91 [0.52;1.58] | 0.74 | 1.03 [0.62;1.72] | 0.91 |
| **Already 1 case in the household** | **No** | 412 (86.9) | 62 (13.1) | 1 |  | 1 |  |
|  | **Yes** | 688 (80.7) | 165 (19.3) | 1.37 [0.96;1.95] | 0.085 | 1.54 [1.08;2.19] | 0.018<sup>§</sup> |
| **Neighborhood** | **Banankabougou** | 454 (82.5) | 96 (17.5) | 1 | 0.98 | Not included |  |
|  | **Faladie** | 229 (82.4) | 49 (17.6) | 0.98 [0.57;1.70] |  |  |  |
|  | **Yirimadio** | 417 (83.6) | 82 (16.4) | 1.03 [0.67;1.59] |  |  |  |

<sup>§</sup>significant test result
\* n=1327
\*\* n=1323 (4 participants showing negative serology with missing ages)

### Knowledge, attitudes, behaviors, practices (KABP)

The KABP score, using the 13 items (false/true/don’t know questions) described in table 5, showed no mean differences according to gender, with respectively mean=7.9 vs 7.6, p=0.065. Attitudes, behaviors and practices measured by age and gender (table 6a and 6b) showed, at first, a high level of denial on COVID-19 disease: a large part believed that COVID-19 was a punishment from God (43.7%), a belief mainly shared by older people (mean=25.1 years) compared to others (mean=21.7 years). Many participants believed that COVID-19 was introduced in Mali by white people (45.3%). Other opinion was less held among participants: almost one-third (30.3%) thought that COVID-19 was a way used by Malian politicians to take money from developed countries. This last opinion was more shared among men than women (33.6% vs 26.2%, p=0.01). A small proportion of participants believed that COVID-19 was due to a spell (14.8%).

**Table 5:**
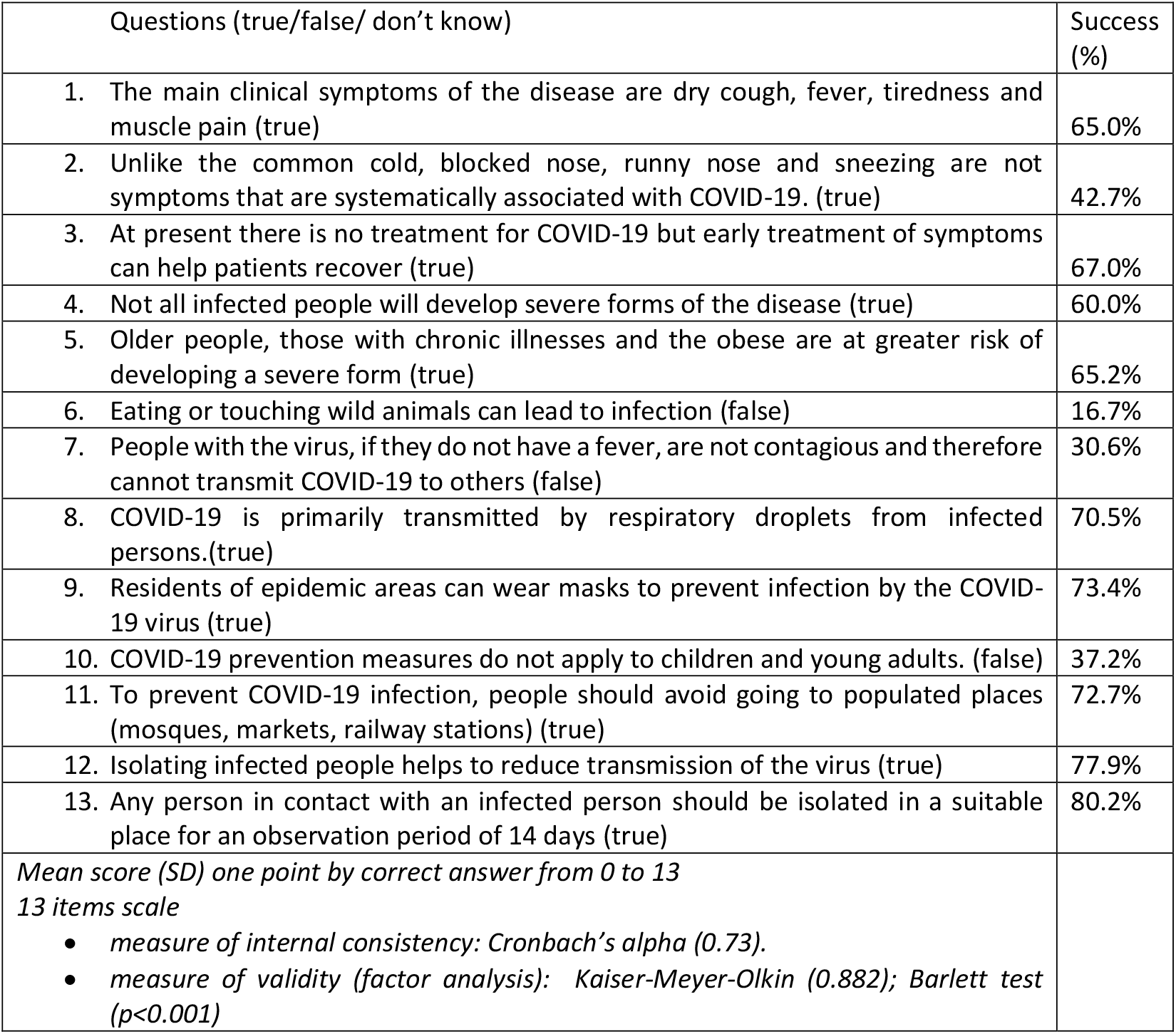
Knowledge on COVID-19 (Bamako, n=962, September 2020).

| Questions (true/false/ don't know) | Success (%) |
| --- | --- |
| 1. The main clinical symptoms of the disease are dry cough, fever, tiredness and muscle pain (true) | 65.0% |
| 2. Unlike the common cold, blocked nose, runny nose and sneezing are not symptoms that are systematically associated with COVID-19. (true) | 42.7% |
| 3. At present there is no treatment for COVID-19 but early treatment of symptoms can help patients recover (true) | 67.0% |
| 4. Not all infected people will develop severe forms of the disease (true) | 60.0% |
| 5. Older people, those with chronic illnesses and the obese are at greater risk of developing a severe form (true) | 65.2% |
| 6. Eating or touching wild animals can lead to infection (false) | 16.7% |
| 7. People with the virus, if they do not have a fever, are not contagious and therefore cannot transmit COVID-19 to others (false) | 30.6% |
| 8. COVID-19 is primarily transmitted by respiratory droplets from infected persons.(true) | 70.5% |
| 9. Residents of epidemic areas can wear masks to prevent infection by the COVID-19 virus (true) | 73.4% |
| 10. COVID-19 prevention measures do not apply to children and young adults. (false) | 37.2% |
| 11. To prevent COVID-19 infection, people should avoid going to populated places (mosques, markets, railway stations) (true) | 72.7% |
| 12. Isolating infected people helps to reduce transmission of the virus (true) | 77.9% |
| 13. Any person in contact with an infected person should be isolated in a suitable place for an observation period of 14 days (true) | 80.2% |
| *Mean score (SD) one point by correct answer from 0 to 13*<br>*13 items scale*<br>*• measure of internal consistency: Cronbach's alpha (0.73).*<br>*• measure of validity (factor analysis): Kaiser-Meyer-Olkin (0.882); Barlett test (p<0.001)* |  |

**Table 6a:** Knowledge, attitudes, behaviors and practices toward COVID-19 among Bamako inhabitants (Bamako, n=962, September 2020)

|  | 12-19 years old |  |  | 20-39 years old |  |  |
| --- | --- | --- | --- | --- | --- | --- |
|  | Men | Women | p value | Men | Women | p value |
| *Attitudes/denials towards COVID-19 measured by agreement (agreed, very agreed) with following opinions:* |  |  |  |  |  |  |
| • Is a god Punishment | 40.1% | 41.7% | 0.702 | 41.2% | 44.6% | 0.591 |
| • Has been introduced in Mali by the white people | 46.4% | 43.6% | 0.487 | 48.5% | 46.4% | 0.749 |
| • Is due to a spell | 14.9% | 15.6% | 0.822 | 11.5% | 14.3% | 0.509 |
| • Help politicians' strategy to take money from developed countries | 33.8% | 27.8% | 0.112 | **37.4%** | **24.1%** | **0.026** |
| *Systematic daily changes in behaviors reported from the start of COVID-19 pandemic:* |  |  |  |  |  |  |
| • Washing hands | 27.5% | 24.6% | 0.420 | 35.9% | 35.7% | 0.979 |
| • Blowing into the elbow | 12.6% | 8.4% | 0.099 | 14.4% | 14.4% | 0.996 |
| • Stop touching other people (systematically) | 12.6% | 15.2% | 0.353 | 14.4% | 16.1% | 0.716 |
| • Traveling less frequently | 8.7% | 9.8% | 0.616 | 14.4% | 13.4% | 0.822 |
| • Avoiding populated places | 7.2% | 9.1% | 0.389 | 13.0% | 11.6% | 0.746 |
| • Avoiding seeing friends | 3.9% | 5.7% | 0.299 | 8.4% | 11.6% | 0.403 |
| *At-risk practices during the seven past days declared:* |  |  |  |  |  |  |
| • Wearing mask outside systematically or very often | 27.5% | 24.2% | 0.361 | 32.8% | 33.0% | 0.972 |
| • Visiting populated public places every day or very often | 31.4% | 27.8% | 0.329 | **24.2%** | **40.2%** | **0.008<sup>s</sup>** |
| • Going out every night or very often | 21.3% | 17.5% | 0.250 | 13.0% | 17.9% | 0.291 |
| • Washing hands when necessary | 59.1% | 57.4% | 0.677 | 51.1% | 56.3% | 0.426 |
| • Staying every day, or very often, more than two hours in a small closed space | 22.8% | 22.4% | 0.926 | 20.5% | 20.7% | 0.959 |
| • Had participated to social events every day or very often | 21.3% | 21.6% | 0.921 | 18.9% | 24.1% | 0.326 |

<sup>s</sup>significant, after Bonferroni correction

**Table 6b:**
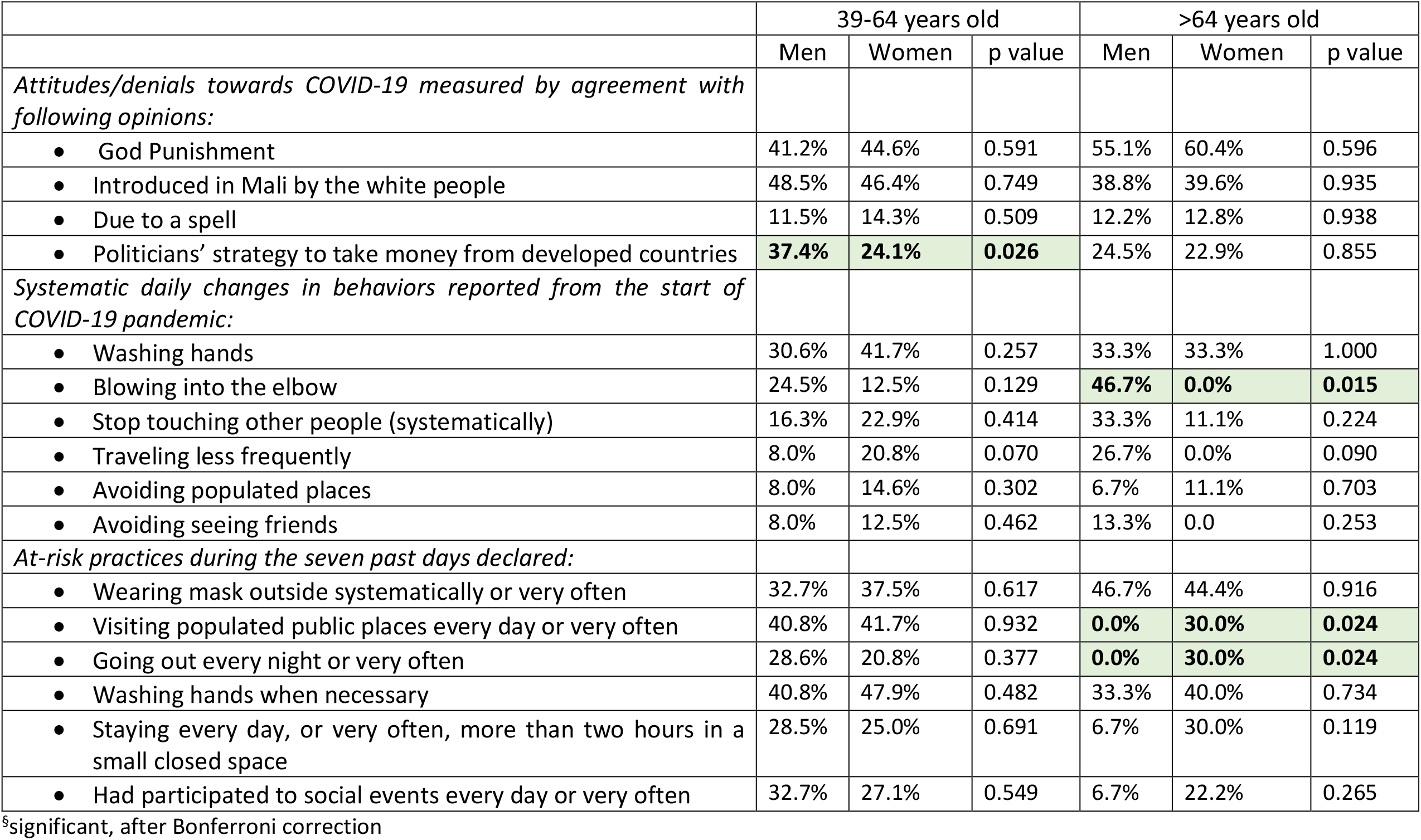
Attitudes, behaviors and practices toward COVID-19 among Bamako inhabitants (Bamako, n=962, 2020)

|  | 39-64 years old |  |  | >64 years old |  |  |
| --- | --- | --- | --- | --- | --- | --- |
|  | Men | Women | p value | Men | Women | p value |
| *Attitudes/denials towards COVID-19 measured by agreement with following opinions:* |  |  |  |  |  |  |
| • God Punishment | 41.2% | 44.6% | 0.591 | 55.1% | 60.4% | 0.596 |
| • Introduced in Mali by the white people | 48.5% | 46.4% | 0.749 | 38.8% | 39.6% | 0.935 |
| • Due to a spell | 11.5% | 14.3% | 0.509 | 12.2% | 12.8% | 0.938 |
| • Politicians' strategy to take money from developed countries | **37.4%** | **24.1%** | **0.026** | 24.5% | 22.9% | 0.855 |
| *Systematic daily changes in behaviors reported from the start of COVID-19 pandemic:* |  |  |  |  |  |  |
| • Washing hands | 30.6% | 41.7% | 0.257 | 33.3% | 33.3% | 1.000 |
| • Blowing into the elbow | 24.5% | 12.5% | 0.129 | **46.7%** | **0.0%** | **0.015** |
| • Stop touching other people (systematically) | 16.3% | 22.9% | 0.414 | 33.3% | 11.1% | 0.224 |
| • Traveling less frequently | 8.0% | 20.8% | 0.070 | 26.7% | 0.0% | 0.090 |
| • Avoiding populated places | 8.0% | 14.6% | 0.302 | 6.7% | 11.1% | 0.703 |
| • Avoiding seeing friends | 8.0% | 12.5% | 0.462 | 13.3% | 0.0 | 0.253 |
| *At-risk practices during the seven past days declared:* |  |  |  |  |  |  |
| • Wearing mask outside systematically or very often | 32.7% | 37.5% | 0.617 | 46.7% | 44.4% | 0.916 |
| • Visiting populated public places every day or very often | 40.8% | 41.7% | 0.932 | **0.0%** | **30.0%** | **0.024** |
| • Going out every night or very often | 28.6% | 20.8% | 0.377 | **0.0%** | **30.0%** | **0.024** |
| • Washing hands when necessary | 40.8% | 47.9% | 0.482 | 33.3% | 40.0% | 0.734 |
| • Staying every day, or very often, more than two hours in a small closed space | 28.5% | 25.0% | 0.691 | 6.7% | 30.0% | 0.119 |
| • Had participated to social events every day or very often | 32.7% | 27.1% | 0.549 | 6.7% | 22.2% | 0.265 |

<sup>§</sup>significant, after Bonferroni correction

Concerning changes in daily preventive behaviors from the start of the COVID-19 pandemic, hand washing was reported as the most used by people: only 4.9% of the participants declared rarely, very rarely or never washing hands in their daily life, compared to 29.9% who declared washing hands systematically. Conversely, few participants reported adopting systematically other preventive behaviors in their daily life, such as blowing into the elbow (12.9%), stop touching other people (15.0%), traveling less frequently (11.1%), avoid populated places (9.3%), and avoiding seeing friends (6.6%). Regarding results displayed by age and sex (tables 6a and 6b), the youngest participants were more reluctant to change their daily behaviors whatever their gender.

Finally, most of the participants declared having at-risk practices during the 7 last days, such as never wearing mask when outside (32.7%), visiting very often or daily highly populated public places (31.0%), going out very often or every night (26.1%), not washing hands most of the time (43.2%), staying in closed spaces more than two hours daily or very often (22.4%), or participating to social or family events daily or very often (40,3%). Young participants declared wearing mask less systematically or very often: mean age=22 years vs mean age=25 years). Young women also declared more visiting populated public places than men (40.2% vs 24.2%, 20-39 year old)

## Discussion

SARS-CoV-2 population adjusted seroprevalence in the urban commune VI of the Bamako district was 16.4%. This prevalence was much higher than the cumulative incidence reported by epidemiological surveillance since the beginning of the pandemic on the investigation site, which was 0.07% at the time of this survey (September 2020). It can be assumed that there was still active circulation of the virus in the capital city at the time of the surveys, suggested by the presence of IgM positive individuals. The corrected survey data suggest that a high number of SARS-CoV-2 infections occurred in the study site. Projected on the total population of Bamako, this prevalence would correspond to a total of 397,321 cases in September 2020. Mortality projections are crude but suggest that deaths caused by COVID-19 were also under-reported, with 81 reported for an estimated 1,720 expected deaths in Bamako in September 2020. The presence of IgM positive individuals suggest the persistence of active viral circulation at the time of the survey.

Seroprevalence was significantly lower in the Kenyan study, reporting 5.6% in a sample of 3,098 blood donors during the same period [15]. This study found a higher prevalence in urban cities and more widespread circulation of SARS-CoV-2 than reported by case-based surveillance. A similar study conducted in Kinshasa, Democratic Republic of Congo, in October-November 2020 after the first wave found a prevalence of 16.6%, a value close to that estimated here [7]. The differences between the different districts of the Congolese capital were not significant, as in the commune VI of Bamako. In Mali, Sagara *et al*. reported in the peri-urban area of Sotuba a crude seroprevalence of 13.1 % (n=587) across samples collected over a 2-month period after the first wave. But the subsequent study conducted in January 2021 in this peri-urban area showed an adjusted seroprevalence rate of 73.4%, after the second COVID-19 wave [6]. This sharp increase in the prevalence rate can be explained by a wave of intense transmission of COVID-19 related to alpha variant in Mali between November 2020 and January 2021 together with the increase of the screening capacity of the health services [4].. Indeed, 3,258 new cases were officially reported at the Bamako district (and 172 new deaths) between November 1rst 2020 and January 24 2021.

In our study, seropositivity was higher among women, with a predominance in the 20-40 age group. Conversely, in Senegal, a survey of the acceptability of the measures to fight the COVID-19 found a predominance of the 25-59 years age and male group [16]. Similarly, a literature review on seroprevalence among health workers worldwide found a seroprevalence of 8.2% in Africa with a male predominance [17]. This difference may be explained by the methodology of our study, which recruited only in households and during the day, *i*.*e*. working time: men aged 20 to 60 may be under-represented in our sample. We did not find any difference in symptomatology between COVID-19 positive and negative individuals during the last four months of the survey. This confirms the pauci-symptomatic clinical situation of the disease. The main demographic characteristics (age and gender) and proximity as a high potential contact rate (a household member already infected) remained significantly associated with seropositivity after adjusting for the contextual elements available. Although the household condition profile was not a significant determinant of seropositivity, the impact of infection among rich family units should be discussed (aOR 1.74 [0.99;3.07]). Indeed, poor families are more likely to live outdoors, to have lower ages, to have fewer co-morbidities (obesity, diabetes) in this population. The age-related results were consistent with the epidemiological trends observed during the first wave of the epidemic worldwide: young people were less exposed than older one despite the higher level of risk practices, revealed by the KABP survey, and, as a result, were more reluctant to change their health behavior. According to psychological models of preventive behavior, self-perceived exposure is a key component of individual acceptability of preventive behavior change [18]. Nevertheless, hand washing was a common practice, perhaps associated with former epidemic (e.g. Ebola in 2014), but not mask wearing, a little-know health practice in the Malian culture. Conversely, gender differences in outcomes remain problematic. Given the complexity of the relationship between sex, gender, and infectious disease [19], the updated medical literature reports greater vulnerability of men to COVID-19 than women due to gender-related social activities or comorbidities, but also due to significant sexual variations in the immune system [20, 21]. The vulnerability of women highlighted by our survey refers to a broader conception of the impact of SARS-CoV-2, including the carriage of the infection. However, with respect to the KABP survey results, with the exception of a tendency for women to score lower on knowledge of COVID-19, no significant statistical evidence emerged on an association between gender and health behaviors and risk practices. A possible selection bias in the serological survey could partly explain these results, but other hypotheses concerning the specific lifestyle and social position of West African women in light of exposure to infectious diseases need to be further explored. Furthermore, the results of multivariate analyses showing the role played by proximity in person-to-person transmission confirm that the spread of infectious diseases within the community involves a significant amount of within family transmissions due to asymptomatic transmission [22], particularly via children[23]. A study on factors associated with the acceptability of government measures against COVID-19 in Senegal showed a correlation between education level and the proposed measures (inter-regional travel ban, curfew, closure of places of worship and closure of markets). But those with primary education and those with no education were likely to accept of curfews and less likely to accept inter-regional travel bans and the closure of places of worship [16]. Finally, the trend of increasing positivity of the social indicator summarized in household profiles leads us to consider that understanding epidemic dynamics in populated cities involves taking into account the spatial structure of the population [24]. Additional evidence from geographic and socio-economic components [25, 26]), highlight the question of inequalities and individual vulnerability at each stage of the epidemic’s spread: from dissemination including various factors such as household size [27], transmission of infection within the community to the associated societal consequences [28].

## Conclusion

In March 2022, 2 years after the pandemic onset and 4 epidemic waves, 30,398 confirmed cases (725 associated deaths) were officially reported in Mali, 20,115 for the district of Bamako, and 60 health districts (among 75) reported cases. The Commune VI remains the most affected (or the most reporting cases) area with 5,712 reported cases. However, these reported numbers under-estimate the number of infected persons. The following waves involved variants, which were more aggressive and may also have led to a heavier death toll, and the consequences could be evaluated using revised prevalence and variant-adjusted infection fatality ratios. Conducted after the first wave, this study highlights the need for sufficient screening data to design efficient epidemic control strategies. Improving diagnostic capacities as well as awareness of populations, to encourage testing and preventive behaviors, as well as avoiding the spread of false information on the epidemic remain key pillars, not matter the developed or developing setting.

## Data Availability

All data produced in the present study are available upon reasonable request to the authors

https://sesstim.univ-amu.fr

## Acknowledgements and funding

We acknowledge the populations of the Commune VI, Bamako, and all the community leaders.

This study was funded by IRD (French National Research Institute for Sustainable Development); the JEAI DynaSTEC (Spatio-Temporal Dynamics of Epidemics and Environmental Changes research team); the French Embassy in Mali (field data collection) and the Charles Mérieux Foundation, Lyon (laboratory analyses); the NGO Prospective and Cooperation.

The funders had no role in the study design, data collection plan, analysis, decision to publish, or preparation of the manuscript.

## Author contributions

JG and HB conceived and designed the study protocol, helped by JL, IS, MC, BK, IB, AG, OD, AD and MKBD. All the authors validated the study protocol.

MKBD wrote the household and KABP questionnaires, with the help of JL, MC, JG and IS.

MC, AKS and IS organized and supervised the samples and data collections, performed by AK, SS, ZD, CD, IT, ST, HM.

ST and IT were in charge of the information system under the supervision of IS.

JL, MKBD, MC and JG conceived and designed the data analysis. JL, MKBD and MC performed the data management and analysis.

BK and AKS designed and supervised the serological analysis, performed by HM, ES, KC. JL, MKBD, MC and JG wrote the paper, corrected by all authors.

All authors participated to the manuscript and approved its last version.

